# Post-SARS-CoV-2 Onset Myalgic Encephalomyelitis/Chronic Fatigue Syndrome Symptoms in Two Cohort Studies of COVID-19 Recovery

**DOI:** 10.1101/2024.11.08.24316976

**Authors:** Armaan Jamal, Thomas Dalhuisen, Nuria Gallego Márquez, Alisha D. Dziarski, Julian Uy, Samantha N. Walch, Sean A. Thomas, Emily A. Fehrman, Arianna E. Romero, Ashley S. Zelaya, Enam A. Akasreku, Tamilore V. Adeagbo, Elizabeth C. Pasetes, Selin Y. Akbas, Alba M. Azola, Steven G. Deeks, J. Daniel Kelly, Jeffrey N. Martin, Timothy J. Henrich, Alan L. Landay, Michael J. Peluso, Annukka A. R. Antar

## Abstract

**Objective:** To determine how many people with long COVID also meet diagnostic criteria for Myalgic Encephalomyelitis/Chronic Fatigue Syndrome (ME/CFS).

**Methods:** We identified which participants with long COVID also met the Institute of Medicine (IOM) or the 2003 Canadian Consensus Criteria (CCC) for ME/CFS at approximately 6-8 months post-SARS-CoV-2 infection in two cohorts: (1) the JHU COVID Recovery cohort, which enrolled participants within 4 weeks of infection and (2) the Long-term Impact of Infection with Novel Coronavirus (LIINC) cohort, which enriched for participants with long COVID. Neither study administered ME/CFS-specific surveys, so available data elements were mapped onto each ME/CFS diagnostic criteria.

**Results:** Of 97 JHU participants with long COVID, 5 met IOM criteria and 2 met CCC criteria. Of 281 LIINC participants with long COVID, 51 met the IOM criteria and 29 met the CCC criteria. In LIINC, participants with long COVID meeting ME/CFS criteria were more likely to be female and report a greater number of post-COVID symptoms (p<0.001).

**Conclusions:** The co-occurrence of ME/CFS symptoms and long COVID suggests that SARS-CoV-2 is a cause of ME/CFS. ME/CFS-specific measures should be incorporated into studies of post-acute COVID-19 to advance studies of post-SARS-CoV-2 onset ME/CFS.

## Introduction

COVID-19 can have long-term health effects. Multiple studies now demonstrate that a significant proportion of individuals infected with SARS-CoV-2 go on to develop long COVID, an infection-associated chronic condition that “occurs after SARS-CoV-2 infection and is present for at least 3 months as a continuous, relapsing and remitting, or progressive disease state that affects one or more organ systems [1].” As of September 2024, it has been estimated that approximately 18% of all US adults had ever experienced long COVID [2]. Common symptoms include fatigue, post-exertional malaise, cognitive dysfunction, shortness of breath, and palpitations [3], which can have a substantial impact on everyday functioning and quality of life.

Many symptoms of long COVID overlap with those of myalgic encephalomyelitis/chronic fatigue syndrome (ME/CFS) [4], a chronic condition characterized by significant impairment in activities compared to the pre-illness time period, profound fatigue, unrefreshing sleep, brain fog, orthostatic intolerance, and worsening of symptoms after physical or mental exertion, among other manifestations [5]. Many symptoms are common to both conditions (e.g., fatigue, post-exertional malaise, headaches, sleep disorder, impaired cognition, myalgia, arthralgia, palpitations, orthostatic intolerance, breathlessness, chills, hot and cold spells, and secondary depression or anxiety) [4]. Others, however, are more commonly associated with long COVID (e.g., disordered smell and taste, rash, and hair loss) or ME/CFS (e.g., painful lymph nodes and chemical sensitivities) [4]. Notably, many people with ME/CFS report symptoms of an infection preceding the onset of ME/CFS [6], suggesting that ME/CFS is likely also an infection-associated chronic condition. Indeed, a small study of the immune cell proteome identified similar molecular pathway enrichment in people with long COVID and those with non-SARS-CoV-2-related ME/CFS [7].

Some people with long COVID also meet diagnostic criteria for ME/CFS, yet estimates of the incidence of post-SARS-CoV-2 onset ME/CFS in the first year of long COVID vary. In studies of people with long COVID recruited from long COVID social media communities and long COVID clinics, between 43-59% of people with long COVID also meet diagnostic criteria for ME/CFS [8-13]. However, it is possible that studies recruiting participants who present to a long COVID clinic or join a long COVID social media community are enriched for more debilitating disease and thus likely to overestimate the incidence of post-SARS-CoV-2 onset ME/CFS compared to studies that followed participants recruited prospectively during acute infection.

In this study, we investigated the proportion of people with long COVID who also meet ME/CFS diagnostic criteria in two observational cohorts of post-COVID recovery, one that enriched for people with long COVID and one that did not. Although we identified an overall lower proportion of individuals meeting ME/CFS criteria in comparison to prior reports, our results demonstrate that post-SARS-CoV-2 onset ME/CFS remains a clinically important phenotype of long COVID warranting additional clinical and biological assessment with dedicated instruments in future studies.

## Methods

### Cohorts

We assessed data from two observational cohorts: the Johns Hopkins University (JHU) COVID Recovery cohort and the University of California, San Francisco (UCSF)-based Long-term Impact of Infection with Novel Coronavirus (LIINC) program.

The JHU COVID Recovery cohort is a prospective, home-based observational study that enrolled participants living in the contiguous 48 United States within 4 weeks of their first known SARS-CoV-2 infection [14]. At enrolment, participants completed a comprehensive symptom survey asking them to recall the presence and severity (Likert scale of 1-5) of 49 long COVID-associated symptoms in the month prior to COVID-19, with an opportunity to write in symptoms not present in the survey. Details of their COVID-19 diagnosis, illness, and treatment history, as well as medical comorbidities, were also collected. Validated surveys such as the Fatigue Severity Scale, the Insomnia Severity Index, the modified Medical Research Council (mMRC) Dyspnea Scale, the General Practitioner assessment of Cognition (GPCOG) self-reported Cognition, the Brief Resiliency Scale, the Breathless, Cough, and Sputum Scale, PHQ-8 (depression), GAD-7 (anxiety), EQ-5D-5L (quality of life) and the Short Form-36 (quality of life), were also administered. Thereafter, comprehensive symptom surveys and neurocognitive assessments were prospectively administered 1, 2, 4, 6, and 12 months post-COVID via telephone or secure web survey [14]. Participant weight and height were measured and biospecimens were collected at 1- and 4-months post-infection. In these surveys, participants were asked about the presence and severity (Likert scale of 1-5) of the same 49 symptoms in the previous week, and they were given the opportunity to write in symptoms as well. The JHU COVID Recovery cohort surveys did not ask the participant to report on whether they thought the symptoms were related to long COVID or not. This study enriched for people living with HIV, with approximately one third of participants reporting an HIV diagnosis. People in the HIV-negative arm had HIV testing performed on blood samples to confirm their HIV-negative status.

As described in detail previously [15], LIINC is an observational study that enrolls individuals with prior test-confirmed SARS-CoV-2 infection in the San Francisco Bay Area. Participants were recruited through self-referrals from general advertisements at a medical center and could enroll any time after 2 weeks following SARS-CoV-2 infection. Participants were offered monthly visits until 4 months post-COVID; thereafter, they were seen approximately every 4 months. Because participants can enter LIINC at any time after SARS-CoV-2 infection, the study is likely to be enriched for individuals with long COVID, even though any individual with test-confirmed COVID-19 is eligible. Detailed clinical interviews at study visits included demographic information, COVID-19 diagnosis, illness, and treatment history, assessment of medical comorbidities and concomitant medications, measurement of height and weight, and evaluation of ongoing symptoms. LIINC surveys evaluated the presence and severity of 34 symptoms within the past two days; participants were also asked about other symptoms not specifically queried. All had to be new, worsened, or persistent since acute COVID-19; symptoms that predated COVID-19 were not considered to represent long COVID. Biospecimens were collected and stored at each study event.

### Ethics approval

Participants in each cohort provided written informed consent. The JHU COVID Recovery cohort was approved by the Johns Hopkins University School of Medicine Institutional Review Board (IRB00278774) and the UCSF LIINC program was approved by the UCSF Institutional Review Board (IRB# 20–30479).

### long COVID definition

We identified participants who met each cohort’s research definition of long COVID at least once between five and twelve months after symptom onset. In the JHU COVID Recovery cohort, a participant was classified as ever having long COVID if, at a survey given at 4, 6, or 12 months post-infection, the participant answered the question ‘Were you at your usual (pre-COVID) health status in the past week?’ as ‘No’ and also reported at least one symptom. For this report, in the JHU study, a participant was also classified as ever having long COVID if the participant reported a higher severity of any symptom at 4, 6, or 12 months post-infection compared to the month prior to COVID-19.

In LIINC, a participant was classified as ever having long COVID if they reported the presence of at least one or more COVID-related symptom at the timepoint closest to 8 months (240 days) post-COVID.

### ME/CFS data elements

Neither the JHU COVID Recovery study nor the LIINC cohort administered ME/CFS-specific surveys, so available data elements were mapped onto the 2003 Canadian Consensus Criteria (CCC) [16] and the 2015 Institute of Medicine (IOM) ME/CFS criteria [17]. Table 1 presents the ME/CFS data elements that were present and not present in each study. The JHU COVID study collected information on 25 of 36 data elements and LIINC did so for 24 of 36. One notable omission was a dedicated question about post-exertional malaise in LIINC, which has since been added to case report forms. Post-exertional malaise was assessed in the JHU cohort via an item in the Fatigue Severity Scale: “During the past week, I have found that exercise brings on my fatigue.”

**Table 1.** ME/CFS data elements queried in each cohort study.

| **Data Elements in both IOM & CCC Diagnostic Criteria** | **JHU** | **LIINC** |
| --- | --- | --- |
| Sleep dysfunction | ✓ | ✓ |
| Cognitive impairment | ✓ | ✓ |
| Fatigue | ✓ | ✓ |
| Difficulty with information processing | ✓ | ✓ |
| Disorientation | ✓ | ✓ |
| General malaise | ✓ | ✓ |
| Reduction in ability to engage in pre-illness levels of activity | ✓ |  |
| Post-exertional malaise | ✓ |  |
| Orthostatic intolerance | ✓ | ✓ |
| Confusion | ✓ | ✓ |
| Light headedness | ✓ | ✓ |
| Extreme pallor |  |  |
| Cold extremities |  |  |
| **Data Elements in only CCC Diagnostic Criteria** |  |  |
| Pain | ✓ | ✓ |
| Impairment of concentration and short-term memory | ✓ | ✓ |
| Nausea | ✓ | ✓ |
| Heart palpitations | ✓ | ✓ |
| Exertional dyspnea | ✓ | ✓ |
| Recurrent fever | ✓ | ✓ |
| Recurrent sore throat | ✓ | ✓ |
| Recurrent flu-like symptoms | ✓ | ✓ |
| Perceptual and sensory disturbances | ✓ | ✓ |
| Irritable bowel syndrome | ✓ | ✓ |
| Headache | ✓ | ✓ |
| Sweating episodes | ✓ |  |
| Worsening stress symptoms | ✓ |  |
| Urinary frequency |  | ✓ |
| Loss of thermostatic stability |  | ✓ |
| Marked weight change |  | ✓ |
| Bladder dysfunction |  |  |
| Loss of adaptability |  |  |
| Tender lymph nodes |  |  |
| Intolerance of extremes of heat and cold |  |  |
| Food sensitivities |  |  |

### Statistical Analysis

Descriptive statistical analysis was performed for all variables. Continuous variables are presented as median (interquartile range, IQR), and dichotomous variables are expressed as absolute frequencies (percentage). The characteristics of ME/CFS patients and non-ME/CFS patients in the LIINC cohort were compared using Wilcoxon rank sum test for continuous variables and Pearson’s χ^2^ tests for categorical variables. A p value of less than 0.05 was considered statistically significant. Statistical analyses were performed using Stata/SE 17.0 (StataCorp, 4905 Lakeway Dr, College Station, TX, USA), R (version 4.4.1) [18], and Microsoft Excel.

## Results

Of 217 post-COVID JHU cohort participants, 97 (45%) met this study’s research definition of long COVID and of 388 post-COVID LIINC participants, 281 (72%) met this study’s research definition of long COVID. The demographics and clinical characteristics of the participants are available in Table 2. The median age at enrolment was 40 in the participants with long COVID in the JHU cohort and 45 in the participants with long COVID in the LIINC cohort. In the JHU cohort, 57% of participants with long COVID were male, and 42% of participants with long COVID were male in the LIINC cohort. While 95% of JHU participants with long COVID were vaccinated prior to their first SARS-CoV-2 infection, only 38% were vaccinated before having COVID-19 in the LIINC cohort. The JHU cohort enriched for people living with HIV and had a higher proportion of individuals with long COVID and HIV (35%) compared to the LIINC cohort (15%). On the other hand, the participants with long COVID in the LIINC cohort had a higher rate of severe acute COVID-19 (15% requiring hospitalization) than those in the JHU cohort (3%). People with long COVID in both cohorts typically reported several post-COVID symptoms (JHU: 8 [IQR: 5, 17], LIINC: 5 [IQR: 2, 9]).

**Table 2.** Characteristics of participants meeting research definition of long COVID in the JHU COVID Recovery cohort and the UCSF LIINC cohort.

| **Characteristic** | **JHU**<br>**Participants**<br>**with Long**<br>**COVID**<br>**(n=97)** | **LIINC**<br>**Participants**<br>**with Long**<br>**COVID**<br>**(n=281)** |
| --- | --- | --- |
| **Age at enrolment, median (IQR)** | 40 (36, 56) | 45 (34, 55) |
| **Sex** |  |  |
| **Male, n (%)** | 55 (57%) | 119 (42%) |
| **Female, n (%)** | 42 (43%) | 162 (58%) |
| **Ethnicity** |  |  |
| **Hispanic, n (%)** | 9 (9.3%) | 61 (22%) |
| **Non-Hispanic, n (%)** | 88 (91%) | 220 (78%) |
| **Race** |  |  |
| **Black/African American, n (%)** | 14 (14%) | 9 (3.3%) |
| **White, n (%)** | 61 (63%) | 167 (61%) |
| **Asian, n (%)** | 14 (14%) | 31 (11%) |
| **BMI, median (IQR)** | 25 (22, 29) | 27 (23, 31) |
| **Vaccination Status** |  |  |
| **1+ vaccines prior to COVID, n (%)** | 92 (95%) | 106 (38%) |
| **Unvaccinated prior to COVID, n (%)** | 5 (5.2%) | 175 (62%) |
| **Acute COVID Severity** |  |  |
| **Required hospitalization, n (%)** | 3 (3%) | 41 (15%) |
| **Did not require hospitalization, n (%)** | 94 (97%) | 239 (85%) |
| **HIV Status** |  |  |
| **People living with HIV, n (%)** | 34 (35%) | 42 (15%) |
| **People living without HIV, n (%)** | 63 (65%) | 293 (85%) |
| **Comorbidities** |  |  |
| **Hypertension, n (%)** | 16 (17%) | 50 (18%) |
| **Diabetes, n (%)** | 2 (2%) | 20 (7%) |
| **Symptom count, median (IQR)\*** | 8 (5, 17) | 5 (2, 9) |
| **Current use (JHU) or history of regular (LIINC) tobacco use** | 8 (8.2%) | 49 (30%) |

\*In the JHU COVID Recovery cohort, each participant was asked to report the presence and severity of 49 symptoms and could also write in symptoms. In the LIINC cohort, each participant was asked to report the presence of 34 new or worsened symptoms.

In the JHU cohort, 5 participants who met the research definition for long COVID also met the IOM definition and 2 people with long COVID also met the CCC definition at 121-232 days post-COVID. The majority of these individuals were living with HIV (4 of the 5 meeting the IOM definition and both who met the CCC definition). In LIINC, 51 participants who met the research definition for long COVID also met the IOM definition and 29 met the CCC definition at a median of 247 days post-COVID. In LIINC, few people with long COVID who met ME/CFS criteria were living with HIV (4 of 51 by IOM, 2 of 29 by CCC).

The characteristics of participants with long COVID who met the IOM and CCC criteria in LIINC are shown in Table 3. Notably, the ME/CFS group had a higher proportion of females than the non-ME/CFS group (75% vs. 46%, p=0.007) and the ME/CFS group had higher median symptom count than the non-ME/CFS group (11 vs. 4, p<0.001). There were no statistically significant between-group differences in age at enrolment, BMI, severity of acute COVID-19, and vaccination status prior to COVID-19. The individuals in the group meeting the CCC definition were also more likely to be female (83% vs. 55%, p=0.004) and more symptomatic (14 vs. 4, p<0.001) compared to the group that did not.

**Table 3.** Characteristics of participants who met the research definition of long COVID who did and did not also meet the IOM and CCC ME/CFS diagnostic criteria in the UCSF LIINC Cohort.

|  | **IOM Definition** |  |  | **CCC Definition** |  |  |
| --- | --- | --- | --- | --- | --- | --- |
|  | **No ME/CFS (n=230)** | **ME/CFS (n=51)** | **p-value<sup>a</sup>** | **No ME/CFS (n=252)** | **ME/CFS (n=29)** | **p-value<sup>a</sup>** |
| **Age, median (IQR)** | 46 (34, 56) | 43 (33,52) | 0.4 | 45 (34, 55) | 46 (36, 57) | 0.6 |
| **Sex** |  |  | 0.007 |  |  | 0.004 |
| **Male, n (%)** | 124 (54%) | 13 (25%) |  | 114 (45%) | 5 (17%) |  |
| **Female, n (%)** | 106 (46%) | 38 (75%) |  | 138 (55%) | 24 (83%) |  |
| **Hospitalized during acute COVID** |  |  | 0.3 |  |  | >0.9 |
| **Hospitalized, n (%)** | 36 (16%) | 5 (10%) |  | 37 (15%) | 4 (14%) |  |
| **Not Hospitalized, n (%)** | 194 (84%) | 45 (90%) |  | 214 (85%) | 25 (86%) |  |
| **Symptom count, median (IQR)** | 4 (2, 7) | 11 (8, 15) | <0.001 | 4 (2, 7) | 14 (12, 16) | <0.001 |
| **BMI, median (IQR)** | 27 (23, 31) | 27 (23, 31) | 0.5 | 26 (23, 31) | 28 (23,31) | 0.5 |
| **Vaccinated for COVID-19** |  |  | 0.13 |  |  | >0.9 |
| **Vaccinated** | 82 (36%) | 24 (47%) |  | 95 (38%) | 11 (38%) |  |
| **Not Vaccinated** | 149 (64%) | 27 (53%) |  | 157 (62%) | 18 (62%) |  |
| **PLWH** | 38 (17%) | 4 (8%) | 0.12 | 40 (16%) | 2 (7%) | 0.3 |
| **Hypertension** | 43 (19%) | 7 (14%) | 0.4 | 47 (19%) | 3 (10%) | 0.3 |
| **History of regular tobacco use** | 45 (20%) | 4 (8%) | 0.05 | 47 (19%) | 2 (7%) | 0.11 |

a. Wilcoxon rank sum test; Pearson's Chi-squared test

## Discussion

Our study of ME/CFS symptoms in two cohorts with different recruitment strategies demonstrates that, although the development of post-SARS-CoV-2 onset ME/CFS may be less common in the era of COVID vaccination as it was in 2020, a clinically significant group of individuals with long COVID exhibit a phenotype consistent with ME/CFS. This suggests that at least some long COVID cases may represent ME/CFS precipitated by SARS-CoV-2 and that the ongoing SARS-CoV-2 pandemic may provide the ability to prospectively study the development of ME/CFS, since infections are a common trigger for this chronic condition. Several overlapping mechanisms, including pathogen persistence, human herpesvirus reactivation, microbiome dysbiosis and microbial translocation, and mitochondrial and metabolic dysfunction have been proposed as drivers of both conditions.

The proportion of individuals with long COVID who also met ME/CFS criteria (2-18%) in our study was considerably lower than that in the published literature to date [8-13]. This is likely due to the liberal research definition of long COVID that was used in these cohorts, the high proportion of individuals vaccinated prior to COVID-19, and the lack of deliberate enrolment of individuals with long COVID in each cohort. Notably, the proportion of LIINC participants with long COVID who met ME/CFS criteria is in line with a study by Tokumasu and colleagues, in which ∼17-18% of people visiting a COVID aftercare clinic for symptoms persisting 4 or more weeks after acute COVID-19 met one or more diagnostic criteria for ME/CFS [19]. However, other studies report percentages ranging from 43-59% [8-13], likely reflecting enrichment for more severe long COVID in dedicated long COVID cohorts and programs. These discrepancies highlight the need for the clinical and research communities to use a single definition of long COVID and highlight the need for rigorous population-based studies to identify the true prevalence of post-SARS-CoV-2 onset ME/CFS.

We identified different proportions of ME/CFS among people with long COVID in the two cohorts (2-5% in JHU versus 10-18% in LIINC). There are several potential explanations for this observation. First, JHU participants were enrolled within 4 weeks of their first SARS-CoV-2 infection, before a person could know whether or not they would experience long COVID. In contrast, participants with persistent symptoms may have been more likely to enroll in the LIINC program. Second, the studies enrolled in different eras of the pandemic. The JHU cohort opened later in the pandemic and predominantly enrolled individuals with breakthrough infections attributed to Omicron variants, while LIINC opened in April 2020 and included many individuals with pre-Omicron variants. Long COVID was more common in the pre-Omicron era [20], potentially contributing to the difference in prevalence between the cohorts. Finally, a higher proportion of people in the JHU cohort were male and were vaccinated prior to first infection, both of which are associated with a lower risk of developing long COVID [20,21].

In LIINC, we also show that those with ME/CFS reported a higher symptom count.

This is similar to another study that compared characteristics of people with and without ME/CFS among those with long COVID [8]. We also found that individuals with long COVID who met the ME/CFS criteria were more likely to be female. Some studies have found a higher proportion of females than males in people with both long COVID and ME/CFS, while others have found similar proportions of males and females with both diagnoses [8,19]. The higher proportion of females among those diagnosed with ME/CFS in this study underscores the potential role of sex-specific factors in both the development and manifestation of these conditions, which is now a priority for the field.

Both cohorts enriched for people living with HIV, providing an opportunity to examine long COVID in this important population. The people with post-SARS-CoV-2 onset ME/CFS in the JHU cohort were mostly people living with HIV, whereas the people with post-SARS-CoV-2 onset ME/CFS in LIINC were mostly not people living with HIV. This likely reflects the greater enrichment for people living with HIV in the JHU cohort, as data from the US suggest that people with HIV are more likely to experience post-acute sequelae than HIV-negative people [22]. It might also suggest that, in the post-Omicron era of widespread immunity to COVID, people with long COVID who go on to develop post-SARS-CoV-2 onset ME/CFS are likely to have predisposing risk factors such as chronic viral infection. However, this cannot be known for sure without dedicated studies in the post-Omicron era.

This analysis has several limitations. First, both studies were developed early in the pandemic and were not specifically designed to assess ME/CFS with validated instruments or clinical assessments. ME/CFS has precise case definitions that require a combination of symptoms for a particular duration of time, generally at least 6 months. Not all elements of the ME/CFS diagnostic criteria were addressed in these research cohorts, and thus we report people with an ME/CFS-like condition without clinical confirmation. Second, both cohorts used inclusive case definitions of long COVID, and it is likely the case that some symptoms attributed to COVID-19 are in fact unrelated. The widespread adoption of a single unified definition of long COVID (e.g., that developed recently by the National Academies [1]), will help us more clearly define the risk of developing post-SARS-CoV-2 onset ME/CFS in individuals with long COVID. Third, differences in the cohort designs precluded us from pooling data into a larger analysis.

In summary, we found that a small proportion of people with long COVID in two research cohorts may be likely to have ME/CFS. Despite its limitations, this study underscores the value of deploying ME/CFS-specific surveys in future post-COVID research studies, even when diagnosis of ME/CFS is not the primary outcome. Our findings also support the argument that there is likely to be value in studying long COVID, post-SARS-CoV-2 onset ME/CFS, and non-SARS-CoV-2-related ME/CFS together, to identify commonalities and differences between the conditions, shared and distinct biological mechanisms, and predisposing host factors. Furthermore, the continued emergence of post-SARS-CoV-2 onset ME/CFS in the setting of the ongoing spread of SARS-CoV-2 means that there is the potential to better understand the development of this condition and how it could be prevented. Ultimately, large, rigorous studies will be needed to more accurately and precisely ascertain the proportion of people with long COVID who meet diagnostic criteria for ME/CFS, and further investigational studies will be needed to determine strategies for prevention and treatment.

## Data Availability

The data sets analyzed for this study are available from the corresponding authors on reasonable request pending IRB approval.

